# Hand Washing Compliance and COVID-19: A Non-Participatory Observational Study among Hospital Visitors

**DOI:** 10.1101/2020.06.02.20120022

**Authors:** Biniyam Sahiledengle, Yohannes Tekalegn, Abulie Takele, Demisu Zenbaba, Zinash Teferu, Alelign Tasew, Tesfaye Assefa, Kebebe Bekele, Abdi Tesemma, Habtamu Gezahegn, Tadesse Awoke, Bruce John Edward Quisido

## Abstract

**Background:** Hand washing remains a key measure for intercepting the dispatch of the Coronavirus disease (COVID-19). However, hand washing must be perpetuated properly using soap and water for at least 20 seconds. In response to the current COVID-19 pandemic, various hospitals have imposed mandatory hand washing to everyone prior entering the facilities, and when leaving. This study aimed to assess the hand washing compliance among visitors of a university referral hospital.

**Methods:** A non-participatory observational study was conducted in the main entrance of the hospital from April 27 to May 3, 2020, to measure hand washing compliance of its visitors. The quality of hand washing was assessed via direct observation for compliance with the recommended World Health Organization (WHO) core steps. Data were collected using Open Data Kit (ODK) mobile application.

**Results:** A total of 1,282 hospital visitors were observed, of which 874(68.2%) were males. Full hand washing compliances were observed among 0.9% (95% CI: 0.4–1.4) of the visitors. Withal, there was no difference in the compliances between genders (0.9% vs 0.7%, *P* = 0.745).

**Conclusion:** Despite the fact that proper hand washing with soap and water is proven to be one of the effective ways in preventing the spread of COVID-19, a significant number of hospital visitors did not practice standard hand washing procedures. Improvements in this measure are urgently needed in the face of the current COVID-19 pandemic.

## Background

Hand washing is reckoned to be one of the paramount measures for preventing the transmissions of Severe Acute Respiratory Syndrome (SARS)-CoV-2 virus, which causes the Coronavirus disease (COVID-19). Hand washing is simple, cost-effective, and one of the first lines of defense in ceasing the spread of the current pandemic. It must be done properly using soap and water for at least 20 seconds [1–3].

Respiratory infection caused by viruses like COVID-19 spread when mucus or droplets containing the virus advances into the body through the eyes, nose, or throat. Most often, this betides through contact with contaminated hands. The risk of COVID-19 transmission is also magnified when people come in contact with an infected person, or when they abut surfaces that an infected person has contaminated [3]. Coronavirus subsist in the hands may be effaced with handwashing using soap and water, and if it’s done properly. If the hands are virtuous, there will be a marginal chance that people can dispatch the virus to the eyes, nose, or mouth of other people, or when they touch their faces [4]. Currently, there is a systematic review apropos hand washing and its risk for respiratory infection, and it implied that hand cleaning can gash the risk of the infection by 16% [5]. It is therefore paramount to wash the hands often and correctly to stop the spread of COVID-19 [1, 2, 6]. The unerring way of hand washing requires five key steps: 1) wet both hands under clean running water; 2) stow a liberal amount of soap to the front and back of the hands, as well as fingertips; 3) rub in between and around the fingers; rub the back of each hand using the palm of the other hand; rub fingertips of each hand to the opposite palm of the other hand, and rub each thumb clasped to the opposite hand for at least 20 seconds; 4) rinse thoroughly under running water, and dry hands with clean paper/towel; 5) turn off the water using a paper towel [1,7,8].

Compliance of the community with the recommended hand hygiene practices has paramount importance in averting the spread of COVID-19. Be that as it may, there are ample attestations that in umpteen settings, hand washing behaviors linger as an area which requisites improvement [1, 9–12]. A recent study by Pogrebna et al found that a country’s hand washing culture is a very compelling predictor of the degree of COVID-19 spread [12].

In response to COVID-19, the World Health Organization (WHO) has strongly recommended that all the state members proffer public hand hygiene stations, and ensuring obligatory usage of these when visitors or personnel enter and leave the facilities, such as hospitals and health centers[2]. Ethiopia reported its first case of COVID-19 on 13^th^ March 2020 [13]. From that time on, the country aggressively engaged and communicated different COVID-19 prevention measures to the general public. As part of its preventive proceedings, the public was notified to regularly wash their hands with soap and water for 20 seconds using the recommended steps. Furthermore, all hospitals should ensure the availability of hand washing stations in their main entrances, and that everyone should wash their hands before entering the facilities and when leaving [14]. Still and all, the acceptance of hospital visitors toward hand washing as a routine habit – and their compliances with the prescribed hand washing techniques were uncertain. In lieu of these, concerns about the poor hand hygiene culture in Ethiopia – and their negative implications in the control of COVID-19 were the motivations behind the undertakings of this study. Accordingly, this study assessed the hand washing compliances among the visitors of Madda Walabu University Goba Referral Hospital, Southeast of Ethiopia.

## Methods

### Study Design and Setting

A non-participatory observation study was conducted for a period of one week (from April 27 to May 3, 2020) to determine the magnitude of hand washing compliance (HWC) of the visitors entering Madda Walabu University Goba Referral Hospital. It is the only referral hospital in Bale Zone, Southeast Ethiopia. Data were collected through direct observations and were executed in the main entrance of the referral hospital for seven consecutive days to appreciate a complete picture of the weekly HWC among its visitors. The observations were imposed in the morning and afternoon (8:30–11:00 A.M and 2:00 –4:30 P.M). These times were chosen because they were the busiest visiting hours. Due to the current COVID-19 pandemic, hand washing facilities and the necessary resources (plain soaps and clean, running waters) were available at the main entrance of the referral hospital. Accordingly, visitors were obliged to wash their hands upon arrival on the hospital premises, and when leaving. Furthermore, large 1.2 m^2^hand washing signs were stationed at the hospital entrance to remind visitors about washing their hands.

### Study Participants and Data Collection Procedures

All visitors who washed their hands before entering and when leaving the referral hospital were observed. Anyone who needs assistance/instruction in washing his/her hands was excluded. The observation was conducted using the recommended WHO hand washing tool [7]. Trained data collectors performed all observations and recorded the data using (Open Data Kit (ODK) Collect mobile Apps. To ensure data quality, the data collectors were informed to observe one visitor at a time. They also made sure that the visitors did not recognize them in order for the observant to display natural handwashing behaviors. Each time, two data collectors were involved, and they stand beside the washing area pretending to wait for someone else, or sometimes, they let their shoes be cleaned at the nearby shoe-cleaning stands.

Prior to the beginning of the study, all data collectors were extensively trained regarding data collection protocols and hand washing steps. To establish the inter-rater reliability, a testing session was imposed to data collectors by simulating hand washing demonstrations by random individuals while they observe them. And the interrater reliability was determined to be 0.86.

As recommended by the WHO, the study recorded and analyzed the recommended hand washing steps: 1) wetting the hands with water, 2) applying enough amount of soap to cover all hand surfaces, 3) rubbing the hand’s palm with the other palm – rotary movement, 4) rubbing the right palm over the left dorsum with interlaced fingers, and vice versa, 5) again palm to palm rubbing with fingers interlaced, back of fingers to opposing palms with fingers interlocked, 6) rotational rubbing of left thumb clasped in the right palm, and vice versa, 7) rotational rubbing, backward and forward with clasped fingers of the right hand in the left palm, and vice versa, 8) rinsing the hands with water, 9) drying them with a clean item/towel, and finally, 10) using a clean item when turning off the faucet [7].

### Operational Definition

#### Full compliance

wetting of hands, covering wet hands with soap, scrubbing all surfaces, including palms, back of the hands, in between fingers, and under the fingernails for about 20 seconds, rinsing well with running water, and finally, dry of hands with a clean cloth or by waving in the air [1].

#### Partial compliance

washing both hands with soap specifically, wetting hands, covering wet hands with soap, washing right palm over left dorsum with interlaced fingers and vice versa, and palm to palm rubbing with fingers interlaced, rinsing well with running water, and finally, drying using a clean cloth for about 20 seconds.

#### Minimal compliance

washing both hands with soap for about 20 seconds, which only includes wetting the hands, covering wet hands with soap, and rinsing well with running water.

#### Non-compliance

rinsing both hands with water only.

### Statistical Analysis

All observed data were collected using ODK and were exported to the Statistical Package for the Social Sciences (SPSS) 20.0 software program for analysis. Based on the operational definitions mentioned, the level of hand washing compliance was computed. Descriptive statistics were computed to summarize the study findings. To test whether hand washing compliance varied across the weekdays and gender, the Chi-square test (χ2) was used. A p-value < 0.05 was considered as statistically significant. Finally, data were presented in tables and graphs, as appropriate.

### Ethical approval and considerations

The study was approved by ethical review committee of Madda Walabu University Goba Referal Hospital Research and Technology Transfer Office and permission was sought from Goba Referral Hospital medical director office (*Ref.no 01/2/12594*). Data collectors were worn protective face masks and kept reasonable physical distance during data collection. The data were collected in a private condition and kept confidential.

## Results

A total of 1,282 visitors were observed, of which 874 (68.2%) were males and 408 (31.8%) were females. Six hundred thirty (47.8%) visitors were observed in the morning schedules (**Table 1**).

**Table 1:** The Characteristics of the Study Participants and their Distributions Over the Weekdays in Goba Referral Hospital, May 2020 (n = 1,282)

| Variable | Category | Frequency | Percent |
| --- | --- | --- | --- |
| Sex | Male | 874 | 68.2 |
|  | Female | 408 | 31.8 |
| Observation Sessions | Afternoon | 669 | 52.2 |
|  | Morning | 613 | 47.8 |
| Days of the Week | Monday | 226 | 17.6 |
|  | Tuesday | 132 | 10.3 |
|  | Wednesday | 231 | 18.0 |
|  | Thursday | 171 | 13.3 |
|  | Friday | 156 | 12.2 |
|  | Saturday | 225 | 17.6 |
|  | Sunday | 141 | 11.0 |

### Hand Washing Compliance

The descriptive analysis of the hand washing steps was shown in **Table 2**. The median (Q1-Q3) duration of hand washing was 22 seconds (14–34). In total, only 0.9% (95%CI: 0.4–1.4) of the study participants were fully compliant with the recommended hand washing techniques. A significant number of hospital visitors did not wash their hands properly or they have washed their hands in less effective ways (minimal, partial, or non-compliant) (**Fig 1**). The areas of the hand most neglected during hand washing among hospital visitors were presented in **Fig 2**.

**Fig 1.**
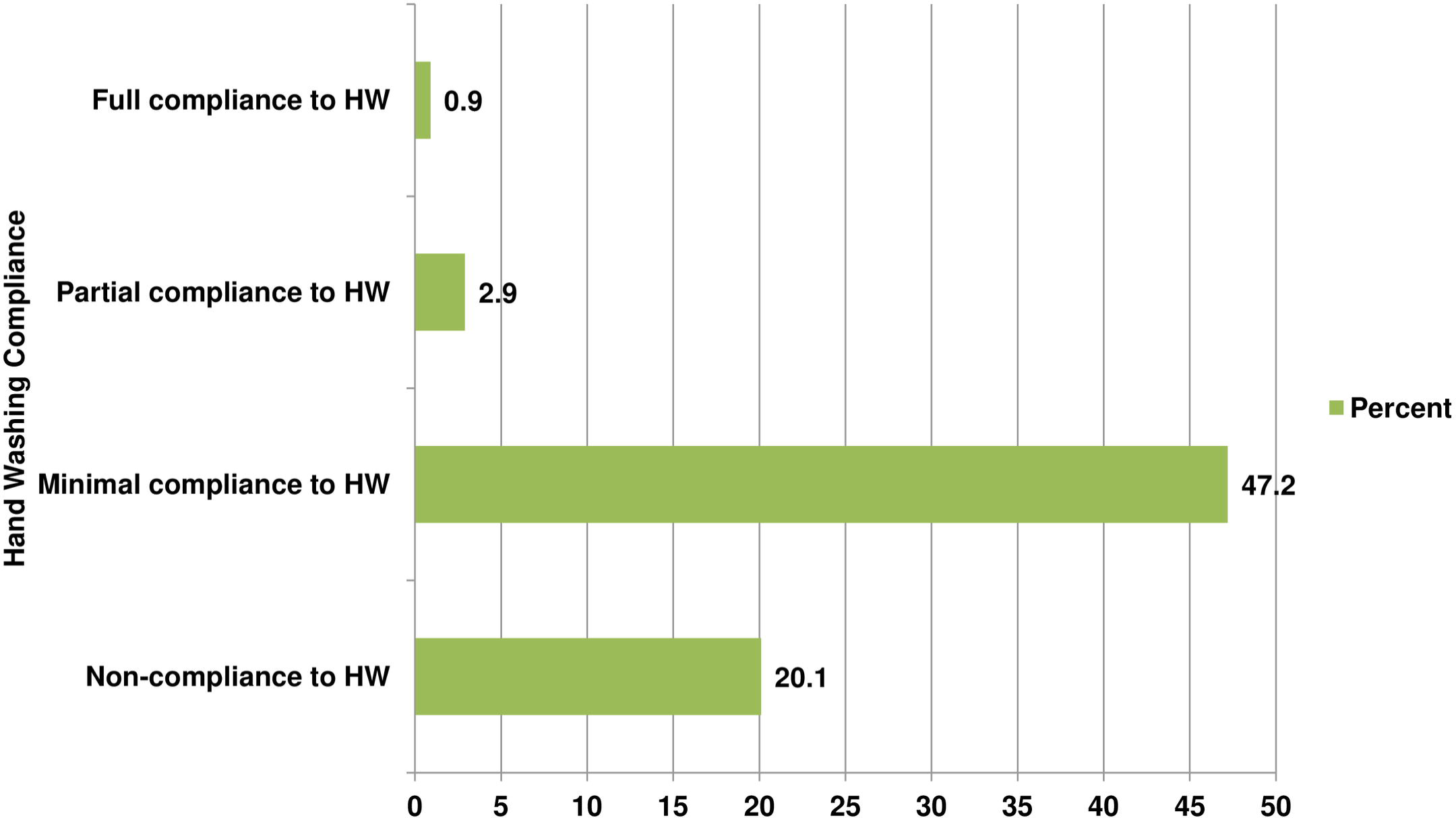
Bar graph including hand washing compliance of hospital visitors at the entrance of Madda Walabu University Goba Referral Hospital, Southeast Ethiopia, May 2020 (n = 1,282)

**Fig 2.**
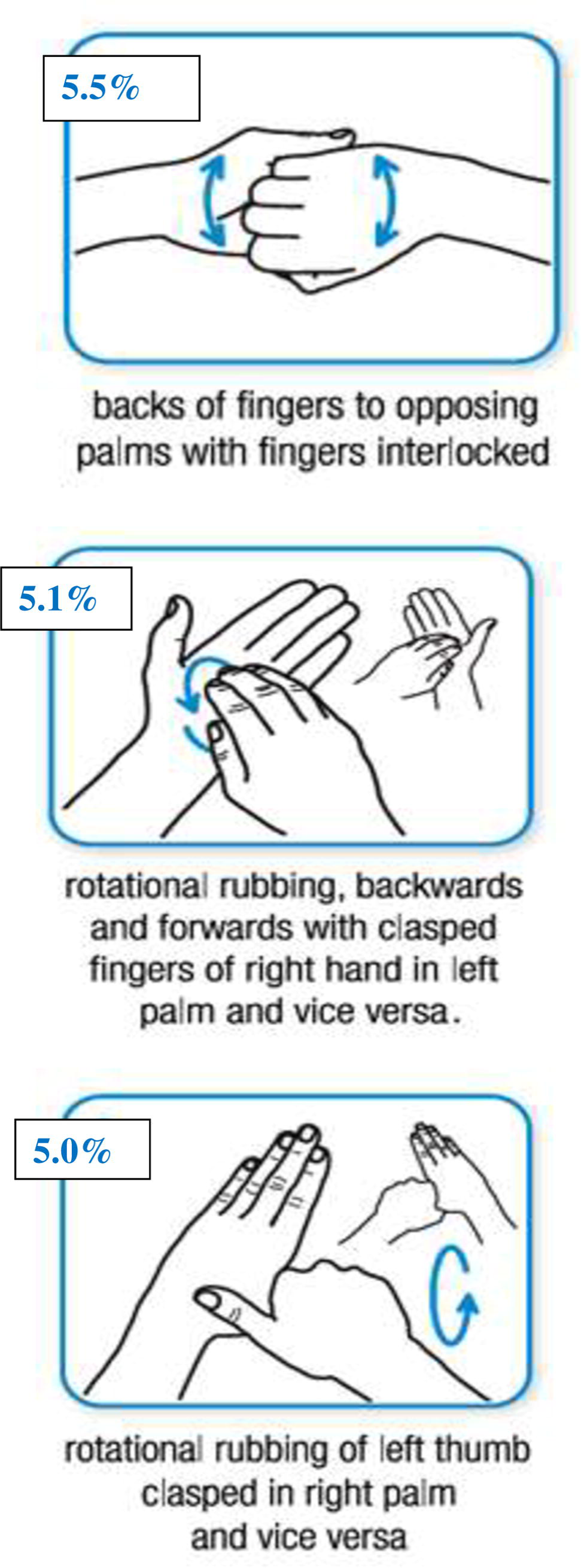
Images of most commonly missed hand washing areas identified among hospital visitors at the entrance of Madda Walabu University Goba Referral Hospital, Southeast Ethiopia, May 2020 (n = 1,282)

**Table 2:**
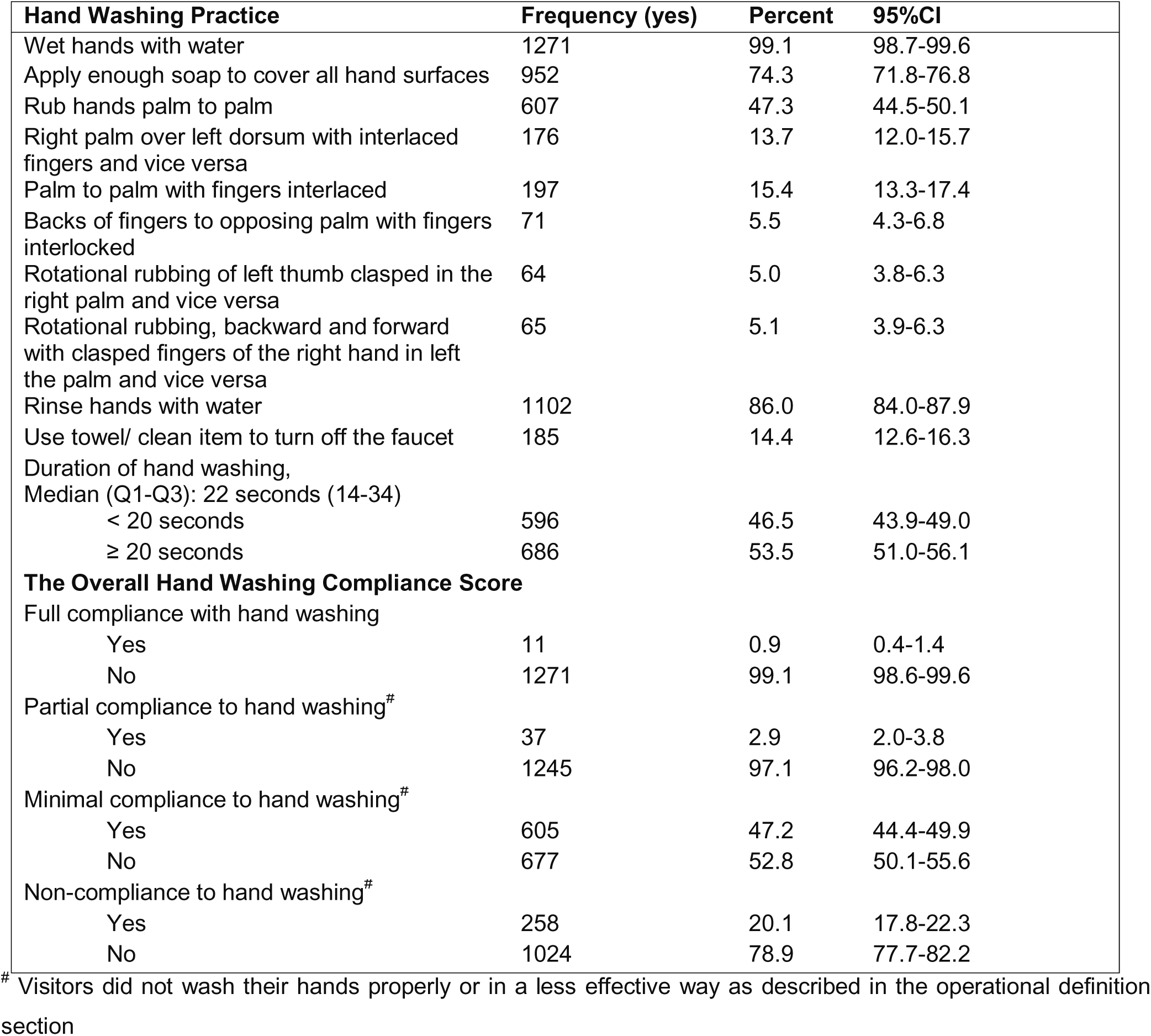
Hand washing compliance in response to COVID-19 pandemic among Study participants of Madda Walabu University Goba Referral Hospital, Southeast Ethiopia (n = 1,282)

| Hand Washing Practice | Frequency (yes) | Percent | 95%CI |
| --- | --- | --- | --- |
| Wet hands with water | 1271 | 99.1 | 98.7-99.6 |
| Apply enough soap to cover all hand surfaces | 952 | 74.3 | 71.8-76.8 |
| Rub hands palm to palm | 607 | 47.3 | 44.5-50.1 |
| Right palm over left dorsum with interlaced fingers and vice versa | 176 | 13.7 | 12.0-15.7 |
| Palm to palm with fingers interlaced | 197 | 15.4 | 13.3-17.4 |
| Backs of fingers to opposing palm with fingers interlocked | 71 | 5.5 | 4.3-6.8 |
| Rotational rubbing of left thumb clasped in the right palm and vice versa | 64 | 5.0 | 3.8-6.3 |
| Rotational rubbing, backward and forward with clasped fingers of the right hand in left the palm and vice versa | 65 | 5.1 | 3.9-6.3 |
| Rinse hands with water | 1102 | 86.0 | 84.0-87.9 |
| Use towel/ clean item to turn off the faucet | 185 | 14.4 | 12.6-16.3 |
| Duration of hand washing,<br>Median (Q1-Q3): 22 seconds (14-34) |  |  |  |
| < 20 seconds | 596 | 46.5 | 43.9-49.0 |
| ≥ 20 seconds | 686 | 53.5 | 51.0-56.1 |
| **The Overall Hand Washing Compliance Score** |  |  |  |
| Full compliance with hand washing |  |  |  |
| Yes | 11 | 0.9 | 0.4-1.4 |
| No | 1271 | 99.1 | 98.6-99.6 |
| Partial compliance to hand washing<sup>#</sup> |  |  |  |
| Yes | 37 | 2.9 | 2.0-3.8 |
| No | 1245 | 97.1 | 96.2-98.0 |
| Minimal compliance to hand washing<sup>#</sup> |  |  |  |
| Yes | 605 | 47.2 | 44.4-49.9 |
| No | 677 | 52.8 | 50.1-55.6 |
| Non-compliance to hand washing<sup>#</sup> |  |  |  |
| Yes | 258 | 20.1 | 17.8-22.3 |
| No | 1024 | 78.9 | 77.7-82.2 |

<sup>#</sup> Visitors did not wash their hands properly or in a less effective way as described in the operational definition section

### Relationship between Hand Washing Compliance and Associated Variables

As shown in **Table 3**, there is no statistically significant difference between male and female hospital visitors concerning hand washing compliance (full and partial compliance). Significant differences were noted in hand washing compliance (full compliance) in between the weekdays (p = 0.03) (**Additional File 2**). No significant differences in hand washing were noted based on time of day (p = 0.875) (**Additional File 2**).

**Table 3:**
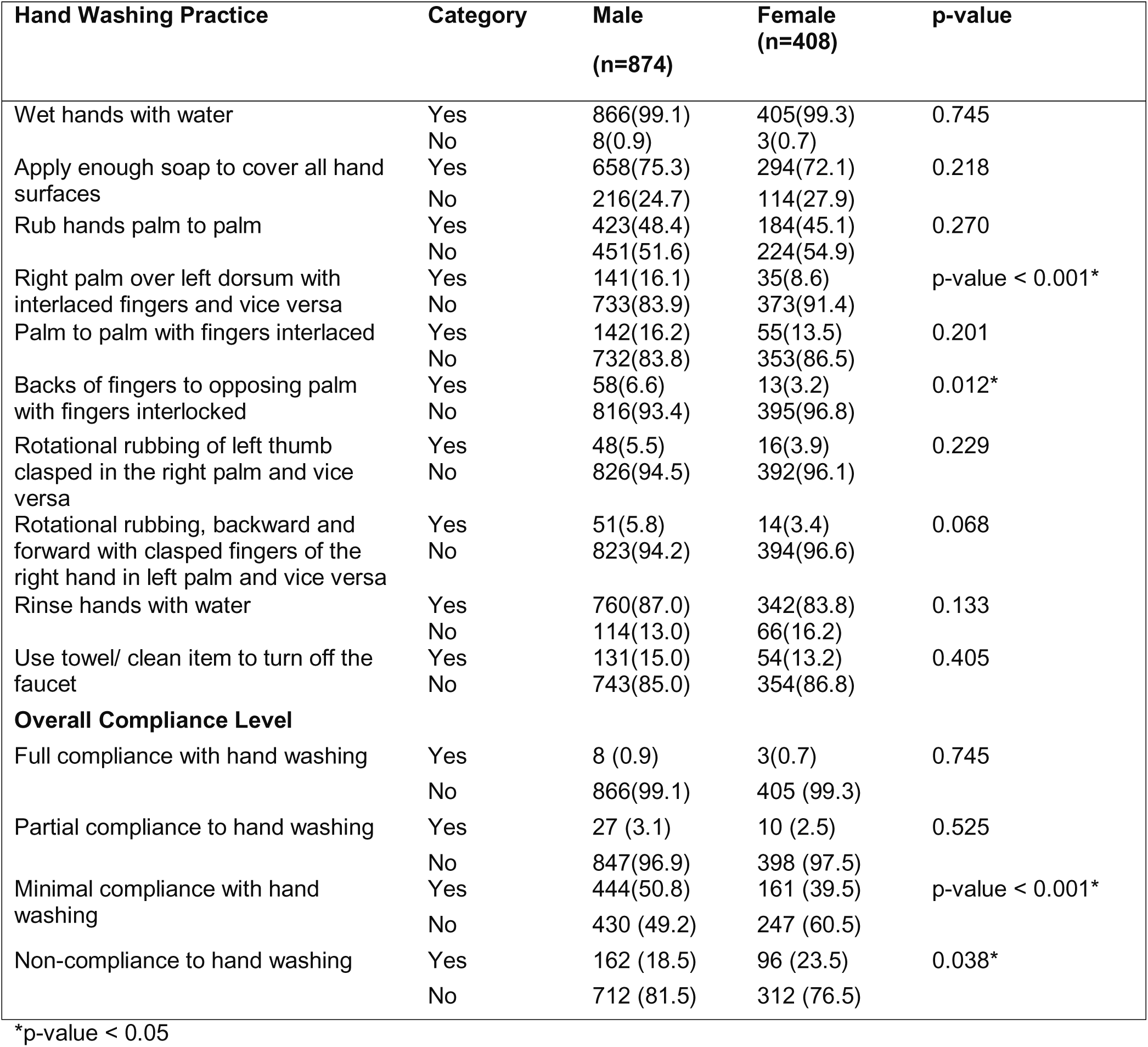
The association between hand washing compliance and sex of the respondents of Madda Walabu University Goba Referral Hospital, Southeast Ethiopia, May 2020 (n = 1,282)

## Discussion

Good hand washing compliance with soap and water for at least 20 seconds is effective in preventing the spread of COVID-19. In response to the current COVID-19 pandemic, the WHO recommended the establishment of hand washing stations located around public areas where people can wash their hands frequently, and somehow prevent themselves from contracting COVID-19 disease. However, public compliance towards the recommended hand washing techniques in these stations is still unknown in many low-income countries, including Ethiopia. Currently, it is now customary to see hand washing facilities in different public areas, yet the question of “Do the people comply with the correct hand washing techniques?” is a fundamental unanswered question. To the best of our knowledge, there is no study done in Sub-Saharan Africa and in Ethiopia, particularly regarding the assessment on the level of hand washing compliance among hospital visitors during this COVID-19 pandemic.

In the current study, only 0.9% of the visitors were observed for full compliance with the recommended hand washing steps. The foremost, very low compliance among hospital visitors is of great concern, and thus highlight the urgent need for interventions from the health authorities, healthcare professionals, and hospital administrators. Also, the findings underscore the need for further behavioral change strategies in increasing the hospital visitors’ compliance, not merely with just the provision of hand washing stations. The level of compliance reported in this study was almost comparable from the baseline assessments in Miami 0.52% [15], and São Paulo, Brazil 1.7% [16]. Despite the similarities between the aforementioned studies, our finding is still extremely low, hence, the present study is conducted during this time of COVID-19 pandemic. Conversely, a relatively higher hand washing adherence by visitors was observed in the United Kingdom 25% [17] and 10.6% [18].

In this study, 46.5% of the study participants wash their hands for less than 20 seconds. According to the CDC, UNICEF, and other experts, a 20-second hand washing is recommended. However, the WHO recommends 40–60 seconds, and this encompasses the entire hand washing process from wetting the hands and applying soap, until the hands are fully dried, whereas, the 20-second recommendation focuses only on the process of scrubbing hands with soap [7, 8, 22, 24]. The challenges behind keeping this duration in public stations need further attention. Some suggest that if people have trouble keeping track, humming the entire “Happy Birthday” song twice before rinsing can help [23]. Failing to meet the proper hand washing techniques and accurate duration can have serious consequences in controlling COVID-19 in counties like Ethiopia, where the healthcare system is not yet matured enough to handle such a pandemic.

Previously conducted studies came up with promising results – audible reminders [19], stylized human watching eye presentations [20], and active methods to encourage hand washing, led to marked improvements on compliance [21]. On the other hand, improving hand washing thru knowledge alone is typically insufficient to change hand washing behaviors [22]. In many cases, humans modify their behavior in a socially desirable way when being observed by others. Hence, applying this basic idea to public hand washing stations may be helpful to positively influence the behavior of individuals [20, 23].

In Ethiopia, hand washing awareness-raising campaigns by some public figures, national radio and television public service announcements, and social media graphics were intensified to raise awareness about the benefits of hand washing in relation to combating COVID-19. Unfortunately, regardless of these efforts mentioned, there are still little improvements in the hand washing habits of the community. Answering the reasons and factors behind the current low compliance is beyond the scope of this study.

There are few limitations in our study. First, we did not use the electronic hand washing observation system which offers a greater advantage over human observations. Second, despite our assumption, the presence of the data collectors may have influenced the hospital visitors’ hand washing activities, and it is likely to have a ‘Hawthorne effect’. Third, selectivity may be possible as the data collectors do not collect data from all people entering and leaving the health facility.

## Conclusion

In conclusion, compliance with proper hand washing among the visitors of Goba Referral Hospital in that specific time period is alarmingly low. A significant number of hospital visitors did not wash their hands properly or washed them in less effective ways. In this era of COVID-19 pandemic, mandating compliance towards proper hand washing to all hospital visitors before entering the hospital and leaving it should be of paramount importance.

## Data Availability

The datasets used and/or analyzed during the current study are available from the corresponding author on reasonable request.

## Acknowledgments

The authors would like to thank Madda Walabu University Goba referral hospital public health department staff for providing their unreserved support. We would like to acknowledge Goba Referral Hospital staff for their support at a time of data collection.

## Supporting Information

**Additional File 1**: The Association between Hand Washing Compliance during the Weekdays in Madda Walabu University Goba Referral Hospital, Southeast Ethiopia, May 2020 (n = 1,282)

**Additional File 2**: The association between hand washing compliance and observation sessions in Madda Walabu University Goba Referral Hospital, Southeast Ethiopia, May 2020 (n = 1,282)

